# Profiling of circulating chromosome 21-encoded microRNAs, miR-155 and Let-7c, in Down Syndrome People

**DOI:** 10.1101/2020.10.24.20218677

**Authors:** Jesús Manuel Pérez-Villareal, Katia Aviña-Padilla, Evangelina Beltrán López, Alma Marlene Guadrón-Llanos, Esther López-Bayghen, Javier Magaña-Gómez, Marco Antonio Meraz-Ríos, Alfredo Varela-Echavarría, Carla Angulo-Rojo

## Abstract

Down syndrome (DS), or Trisomy 21 (Ts21), is the most common chromosomal survival aneuploidy. Nevertheless, people with DS have compromised health, and the increase in their life expectancy further heightens the risk of developing chronic degenerative diseases such as obesity, dyslipidemias and diabetes mellitus associated with higher morbidity, and mortality for cardiovascular disease from an early age. DS is also accompanied by a higher risk of neurodegeneration. The extra genetic material that characterizes DS causes an imbalance in the genetic dosage, including overexpression of miR-155 and Let-7c miRNAs, both associated with cognitive impairment and dementia in adults. The dynamics of expression of their putative target genes in the early stages of the development of DS and their clinical associations, however, remain to be ascertained. This study aimed to evaluate the relative expression of miR-155 and Let-7c in young and adult individuals with DS and its possible association with biochemical indicators of lipid metabolism. The anthropometric, clinical, biochemical, and gene expression features of miR-155 and Let-7c were analyzed in a population of 52 control and 50 DS subjects divided into groups of 20 years of age or younger and 21 years or older. Expression changes for miR-155 were not significant. Nevertheless, a negative correlation for HDL-Cholesterol concentrations and miR-155 expression was identified. Notably, Let-7c was overexpressed in DS from young and old ages. Overall, our results suggest that Let-7c is related from early stages to cognitive impairment in DS, while a similar role of miR-155 in late stages could be mediated by alterations in lipid metabolism. Further studies with both miRNAs will shed light on their potential as therapeutic targets to prevent or delay cognitive impairment in DS.

## Introduction

Down syndrome (DS), or Trisomy 21, is the most common survivable chromosomal aneuploidy, occurring at an approximate rate between 6 and 13 in 10,000 births (1, 2). The prevalence has been increasing in recent decades as the life expectancy of people with DS has expanded to 60 years (3). In addition to cognitive dysfunction, several clinical symptoms have been described for the DS population, including craniofacial abnormalities, gastrointestinal anomalies, congenital heart disease, childhood leukemia, and early-onset Alzheimer’s disease (AD) (4). Moreover, DS individuals have an increased risk of overweight, obesity, and dyslipidemias, which confer a high risk of cardiovascular disease in general, ischemic heart disease, and stroke death compared to the euploid population (5). Obesity in DS is a multifactorial disease mainly associated with high caloric diets, low physical activity, higher serum leptin levels, decreased resting energy expenditure, and other comorbidities such as sleep apnea, hyperinsulinemia, gait disorders, and dyslipidemias (5). Most of the features of DS are expected to be related to the overexpression of the genes located on chromosome 21, particularly those of the Down Syndrome Critical Region (DSCR), leading to imbalanced interactions with other disomic genes (6). Accordingly, microRNA (miRNA) genes located on chromosome 21 could play a role in the DS phenotype, mostly unknown (4).

miRNAs are small non-coding strands of RNA (18–25 nucleotides), which post-transcriptionally regulate almost 60% of all gene expression via inhibition of translation or direct mRNA target degradation. These regulatory RNAs often have tissue-specific expression, are detectable in peripheral blood, and may act as endocrine effectors on target cells. Human chromosome 21 has more than 400 genes, including five miRNAs (miR-99a, Let-7c, miR-125b-2, miR-155, and miR-802) whose expression leads to haploinsufficiency for their mRNA target (7). miRDB database analysis (http://mirdb.org/index.html) (8), predicted that miR-99a is a potential interactor of 47 mRNA targets, Let-7c of 990, miR-125b-2 of 585, miR-155 of 701, and miR-802 of 631. This led us to hypothesize that dysregulation of these miRNAs may play a role in the cognitive dysfunction, congenital heart defects, childhood leukemia, and early-onset Alzheimer’s disease (AD) in individuals with DS.

Among those miRNAs, we focused on Let-7c and miR-155, which are critically related to neurodegeneration and play a crucial role in the DS pathogenesis of dementia (9, 10). AD is the most common form of dementia, and it is characterized by two hallmarks, the amyloid plaques formed by Aβ peptide deposits and the neurofibrillary tangles caused by hyperphosphorylation and accumulation of Tau protein (11). In DS individuals, the increased risk of early-onset Alzheimer disease is related to the trisomic dosage of the genes for amyloid precursor protein (APP) and β secretase 2 (BACE2), essential proteins for Aβ peptide production (12). In addition, the extra copies of miR-155 and Let-7c may contribute to AD, as miR-155 is related to NF-kB signaling and T cell activation and infiltration in the hippocampal parenchyma of the brain with AD, contributing to neuroinflammation (9). Also, miR-155 is co-expressed with hyperphosphorylated Tau (9, 12), and Let-7c overexpression leads to morphological as well as functional deficits, including impaired neuronal morphologic development, synapse formation, and synaptic strength, as well as a marked reduction of neuronal excitability (10, 13). This miRNA dysregulation has also been linked to impaired mitochondrial function (14), neurodegeneration (15), and neurodevelopmental disorders (10). while DS and AD have been found to have robust associations to dyslipidemias, particularly low HDL-Col concentration, common conditions in DS patients (16-19).

Therefore, in the present work, we focused on the study of the expression profiles of miR-155 and Let-7c miRNAs in DS and determining their association with biochemical alterations related to lipid metabolism.

## Materials and methods

### Study design

A cross-sectional study was performed with 50 participants with DS and 52 healthy controls. Individuals were selected at Down Syndrome Health Fairs organized by the Autonomous University of Sinaloa. The study followed the provisions of the Declaration of Helsinki and Good Clinical Practice guidelines, and the local institutional review board approved the protocol. The research project was reviewed and approved by the Research Ethics Committee at the Medicine Faculty of Autonomous University of Sinaloa, which has a national certification (CONBIOÉTICA-25-CEI-003-20181012) for its duties. Parents or legal guardians of people with DS were informed before the evaluation and provided written consent to undergo study procedures.

DS individuals and controls were each grouped in 20-year old or younger (Group A) and 21-year-old or older (Group B), according to the growth standards for children with DS by the Disease Control and Prevention Center (CDC).

### Patients

The inclusion criteria for this study were age over eight years and a DS diagnosis by clinical and genetic criteria. Sex and age-matched control participants were used as a reference. Exclusion criteria were the presence of congenital heart disease not treated with surgery, severe sensory impairment, leukemia, and diagnosis of diabetes mellitus or metabolic syndrome.

### Anthropometric measurement

Anthropometric measurements were carried out following the procedures of the International Society for the Advancement of Kinanthropometry (ISAK) during patient examinations. Bodyweight was measured on subjects wearing light clothes using an electronic bascule (Tanita HS-302) and height using a stadiometer (SECA 213). Body mass index (BMI) was calculated, and individuals were classified according to the BMI/Age tables of the World Health Organization (WHO).

### Biochemical analysis

After overnight fast blood samples were obtained from an antecubital vein. Blood was collected into two non-additive tubes, which were incubated 30 min at room temperature and spun at 3000 × *g* at 4°C for 10 min. Serum samples were collected and stored at –80°C. Serum lipids (total cholesterol and HDL-cholesterol), triglycerides, and plasma glucose were measured using commercial enzymatic kits (HUMAN Diagnostics Worldwide; Wiesbaden, Germany). LDL-cholesterol was calculated using a modified Friedewald equation (20). For the classification of biochemical parameters, we used the cut-off points of the International Diabetes Federation (IDF) and the National Cholesterol Education Program (NCEP) for children, adolescents, and adults (21, 22).

### RNA isolation

Total miRNA purification was performed with the miRNeasy-serum/plasma extraction kit (Qiagen, Hilden, Germany) according to the manufacturer’s protocol. RNA extracted was quantified using the NanoDrop 2000 spectrophotometer (Thermo Fisher Scientific, MA, USA) measuring absorbance at 260 nm and 280 nm. Samples were eluted with RNase-free water for subsequent cDNA synthesis and stored at -80°C.

### Reverse Transcription (RT)

Total RNA was reverse transcribed using a miScript II RT Kit (Qiagen, Hilden, Germany) in a StepOne Plus (Thermo Fisher Scientific, MA, USA) according to the manufacturer’s protocol.

### Real-Time PCR

Quantification of Let-7c (Cat. No. 00003129), and miR-155 (Cat. No. 00031486), mature miRNAs were determined using the miScript SYBR Green PCR kit (Qiagen, Hilden, Germany) and SNORD68 (Cat. No. 00033712) as normalizer. PCR reactions were each performed in a final volume of 15 µl in the StepOne Plus thermocycler. The Melting Curve was performed to observe the specificity of reactions. Relative quantification was obtained using the Pfaffl method, which describes the use of reaction efficiencies obtained using LinReg PCR software (23).

### In silico prediction of miRNAs and their target genes

For the prediction of miR-155 and Let-7c gene targets, miRDB (http://www.mirdb.org) and TargetScan 7.2 (www.targetscan.org) public databases were used (8, 24). To determine the biological processes under each miRNA regulation, we performed network enrichment analysis using Metascape (25) (http://metascape.org/). Then, we used the Meta-analysis workflow to compare miR155 and Let-7c targets and to identify unique and shared biological pathways in which they are involved.

### Statistical analysis

Statistical tests were performed using SPSS v.22. Data were examined for normality based on skewness and kurtosis before analysis. Descriptive statistics of clinical characteristics between groups were represented as means, ± standard deviation. Mann-Whitney U and Student’s t-test were used to examine the statistical difference in clinical parameters between healthy controls subjects and patients with DS. ANOVA Kruskal-Wallis test for differences between group respect to miRNA expression was assessed. Correlations between fold changes of miRNA levels and changes in biochemical characteristics were determined by Pearson correlation coefficients. Statistical significance was set at *p*< 0.05.

## Results

### Anthropometry and biochemical parameters

A total of 50 individuals with Down syndrome and 52 healthy controls from 8 to 52 years of age were characterized and divided into two groups: A (A Control and A DS) and B (B Control and B DS). All individuals were matched by sex and age. The mean age was 10.9±2.4 and 11.3±2.7 years for A Control and A DS groups, respectively; and 28.3±11 and 28.5±11 years for B Control and B DS groups, respectively, with no significant difference between them, which indicates homogeneity of age divided groups. For bodyweight, there was no significant difference between A groups (Control, 41.4±12.5 and DS, 37.9±13.2 kg,) and B groups (Control, 63.3±10.3 kg and DS, 64.3±12.7 kg) (all results are presented in this order heretofore). Nevertheless, we observed a significant difference in height between A groups (1.45±0.11m and 1.3±0.12m, *p=*0.001), and B groups (1.6±0.07m and 1.5±0.08m, *p*=0.001). Moreover, a significant difference in BMI was found between B groups (22.85±2.61 and 28.79±6.03m/kg^2^, p=0.006), while no difference was found between A groups (Table 1). Altogether, these results indicate that the risk of overweight in DS individuals is higher at older stages, in keeping with previous reports (5, 26).

**Table 1.** Participant characteristics and biochemical parameters.

| Clinical | Control A | DS A | *P* value | Control B | DS B | *P* |
| --- | --- | --- | --- | --- | --- | --- |
| Characteristics |  |  |  |  |  | value |
| Age (years) | 10.9±2.4 | 11.3±2.7 | NS | 28.3±11 | 28.5±11 | NS |
| Body weight (kg) | 41.4±12.5 | 37.9±13.2 | NS | 63.3±10.3 | 64.3±12.7 | NS |
| Height (m) | 1.45±0.11 | 1.3±0.12 | **.0001** | 1.6±0.07 | 1.5±0.08 | **.0001** |
| BMI (kg/m<sup>2</sup>) | 19.2±3.3 | 20.5±4.3 | NS | 22.9±2.6 | 28.8±6.0 | **.006** |
| Glucose (mg/dL) | 86.4±11.1 | 87.4±10.4 | NS | 91.08±5.1 | 91±13.6 | NS |
| Cholesterol (mg/dL) | 118.7±30.2 | 181.2±44.6 | **.0001** | 157.4±60.6 | 171.7±54.2 | NS |
| HDL (mg/dL) | 56.8±15.4 | 46.7±13.17 | **.003** | 68.8±13.2 | 42.5±9.1 | **.0001** |
| Triglycerides<br>(mg/dL) | 75.6±43.3 | 139.1±44.5 | **.0001** | 80.3±24.5 | 152.7±83.5 | **.011** |
| LDL (mg/dL) | 109.6±43.1 | 106.4±37.6 | NS | 72.8±55.1 | 115±28.07 | **.029** |

Data are presented as mean ± SD; NS, non-significative; BMI, body mass index; HDL, High-density lipoprotein cholesterol; LDL, Low-density lipoprotein cholesterol.

Biochemical analysis revealed a significant difference between A groups in total cholesterol (118.7±30.2 and 81.2±44.65 mg/dL, *p=0*.*0001*), HDL (56.82±15.4 and 46.7±13.17 mg/dL, *p=* 0.003) and triglycerides (75.65±43.33 and 139.1±44.53 mg/dL, p= 0.0001), but not for LDL and glucose levels. Similarly, for B groups we observed differences in HDL (68.8±13.2 and 42.5±9.1 mg/dL, *p=0*.*0001*), triglycerides (80.3±24.5 and 152.7±83.5 mg/dL, *p= 0*.*011*) and LDL (72.8±55.15 and 115±28.07 mg/dL, *p=0*.*029*), but not for total cholesterol and glucose (Table 1). Overall, these results show alterations in the parameters of lipid metabolism due to the genetic background of DS.

### Serum miRNAs expression

Analysis by qRT-PCR of Let-7c expression revealed a 2.2-fold increase in DS relative to the control group (Fig 1A). Moreover, when age was taken into account, in both age groups, Let-7c expression was found to be higher in DS compared to their respective control groups (DS A: 2.3-fold and DS B: 1.8-fold) (Fig 1A). Nevertheless, no differences were detected for miR-155 in both analyses (Fig 1B).

**Fig 1.**
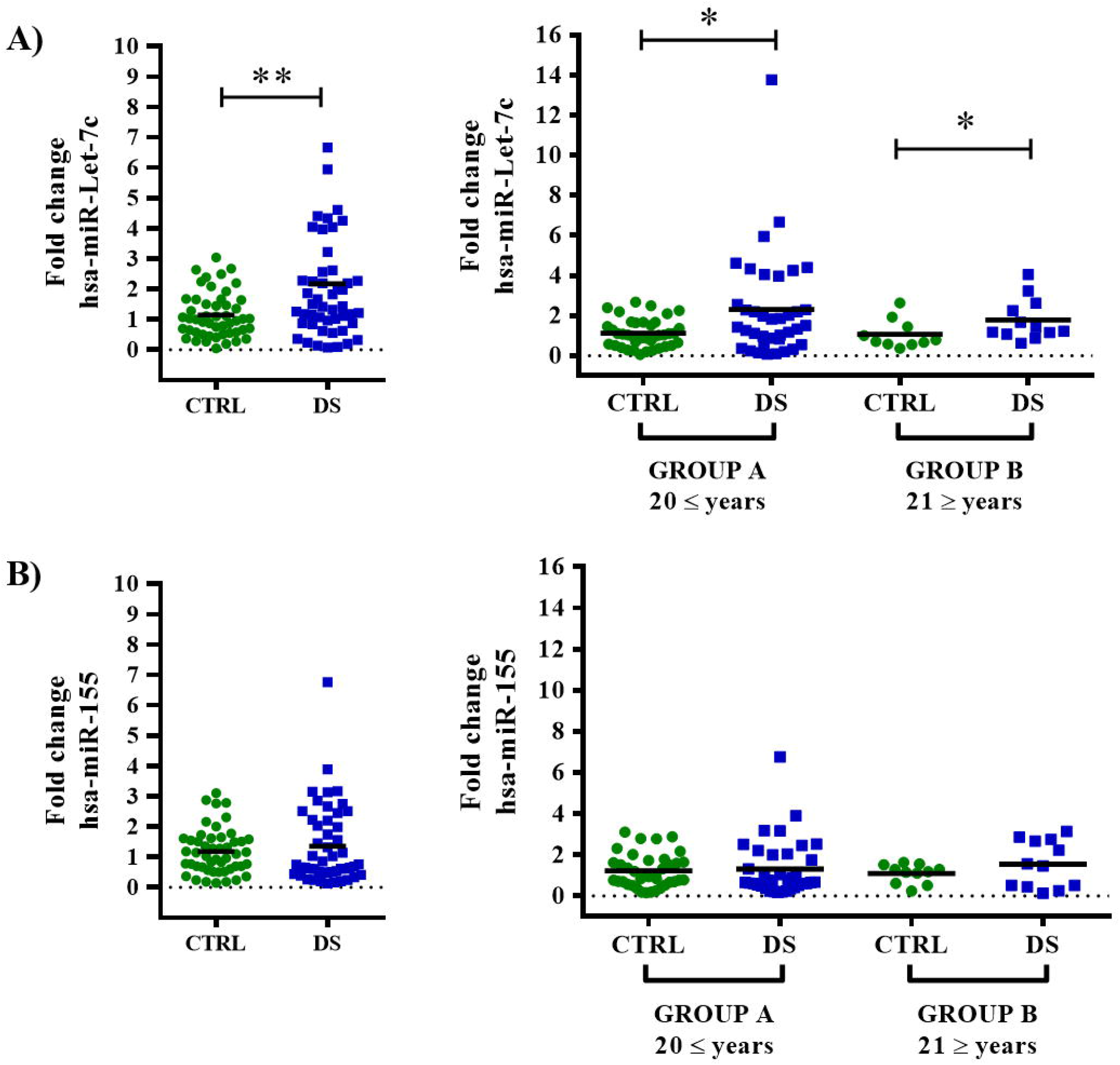
The miRNA Let-7c is up-regulated in DS but not miR-155. Expression of miRNAs Let-7 c **(A)** and miR-155 **(B)** was measured by qRT-PCR in DS people and referenced to control individuals. For each miRNA, total and age-divided population were done. Data are plotted as relative value for each individual, and means for each group statistically analyzed by Mann-Whitney U test (*p<0.05, **p<0.01). DS: down syndrome; Group A: ≤20 years old; Group B: ≥21 years old.

Furthermore, the association of miRNA expression with lipid serum levels was analyzed. For Let-7c, overexpression of 1.7-fold and 2.9-fold was observed when analyzed for Normal-HDL or Low-HDL DS group compared to the control group. These results indicate that DS individuals with lower HDL levels express higher amounts of Let-7c. This behavior was not observed when analyzed the other lipid parameters. A similar analysis performed for miR-155 showed no significant changes (Table 2).

**Table 2.** miRNAs quantification according to lipid concentration, in Down syndrome groups

| miRNAs | DS Normal-HDL | DS Low-HDL | P-value |
| --- | --- | --- | --- |
| Let-7c | 1.7 | 2.9 | .0009 |
| miR-155 | 1 | 1.9 | NS |
|  | DS Normal-TG | DS High-TG |  |
| Let-7c | 2.4 | 1.6 | .0007 |
| miR-155 | 1.4 | 1.2 | NS |
|  | DS Normal-Cholesterol | DS High-Cholesterol |  |
| Let-7c | 2.2 | 2.0 | .02 |
| miR-155 | 1.3 | 1.3 | NS |
|  | DS Normal-LDL | DS High-LDL |  |
| Let-7c | 2.3 | 1.8 | .005 |
| miR-155 | 1.3 | 1.3 | NS |

Data are presented as mean; NS, non-significative; TG, triglycerides; HDL, High-density lipoprotein cholesterol; LDL, Low-density lipoprotein cholesterol.

### Correlation between miRNAs expression and biochemical parameters

Since lipid concentrations have been linked to cognitive impairment, biochemical parameters measured in patients were correlated with expression values of miRNAs Let-7c and miR-155 (Table 3). No correlation was detected for Let-7c levels with any lipid or glucose parameter (Table 3). However, we observed a negative correlation between the HDL levels and the increase in the expression of miR-155 (p <0.003, r = -0.421) (Table 3). Interestingly, an overall landscape showed that almost all of DS individuals have Let7-c miRNA expression up-regulated, and miR-155 expression appears to be up-regulated when HDL levels are far below normal (Tables 2 and 3).

**Table 3.**
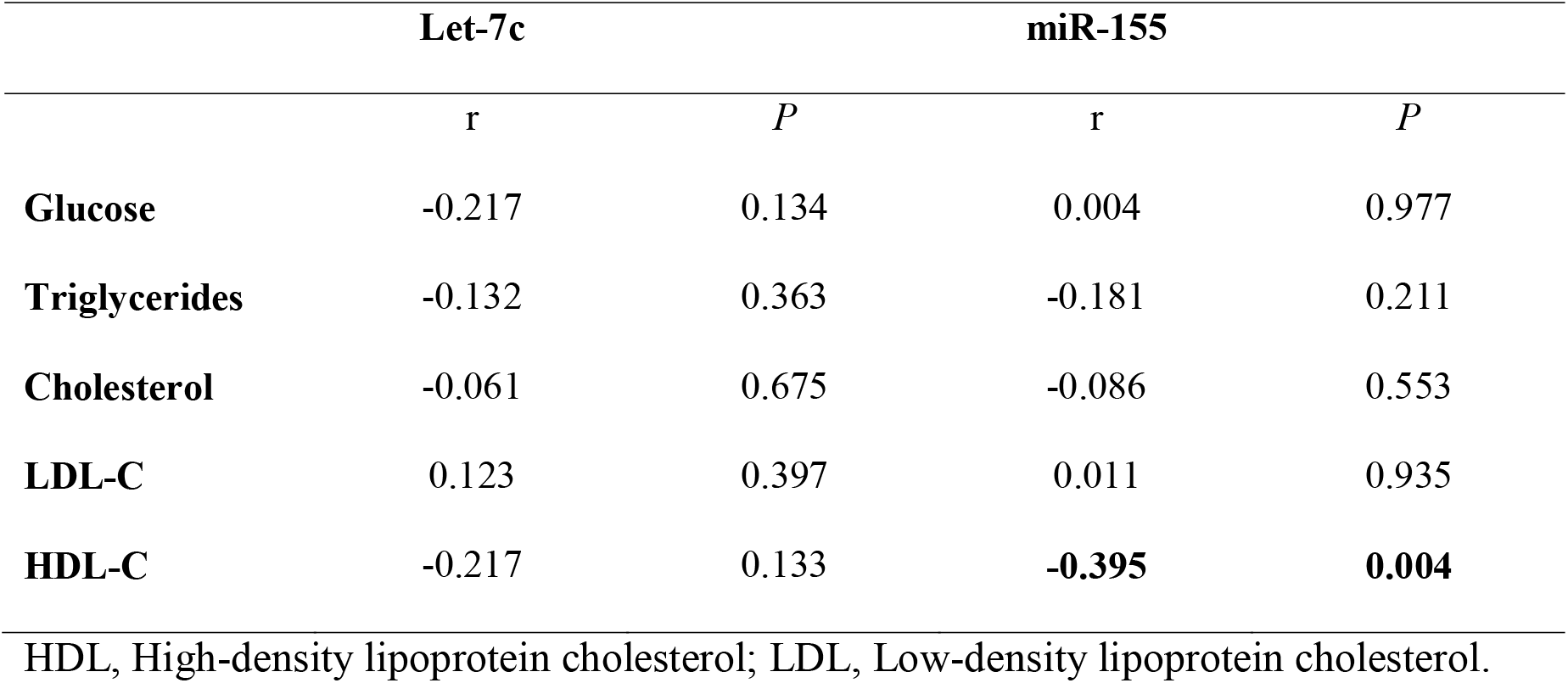
Bivariate correlation of Let-7c and miR-155 levels with changes in clinical characteristics

|  | **Let-7c** |  | **miR-155** |  |
| --- | --- | --- | --- | --- |
|  | *r* | *P* | *r* | *P* |
| **Glucose** | -0.217 | 0.134 | 0.004 | 0.977 |
| **Triglycerides** | -0.132 | 0.363 | -0.181 | 0.211 |
| **Cholesterol** | -0.061 | 0.675 | -0.086 | 0.553 |
| **LDL-C** | 0.123 | 0.397 | 0.011 | 0.935 |
| **HDL-C** | -0.217 | 0.133 | **-0.395** | **0.004** |

HDL, High-density lipoprotein cholesterol; LDL, Low-density lipoprotein cholesterol.

### Targets of miR-155 and Let-7c: unique and shared biological pathways

To gain insight into the targets of miR-155 and Let-7c in DS patients, an *in-silico* analysis was performed to find biological pathways that could be differentially affected. miR-155 and Let-7c targets were determined using the miRDB (http://www.mirdb.org) and TargetScan 7.2 (www.targetscan.org) public databases. Clustering networks of the top 20 enriched GO: BP (Gene Ontology: Biological Processes) terms for miR-155 and Let-7c targets are shown in Fig 2. *In silico* prediction, analysis showed that miR-155 has 701 mRNA targets, while Let-7c could be regulating 990 putative genes. Comparative analysis of their targets showed that only 52 genes could be regulated by both miRNAs (Fig 3 and S1 Fig). Despite the discrepancy in genetic regulation, both miRNAs are linked at the biological pathway level (S1 Fig). It is noteworthy that enrichment analysis of biological processes revealed that the highest p-value enriched GO: BP terms are unique pathways for each miRNA (Figs 2 and 3).

**Fig 2.**
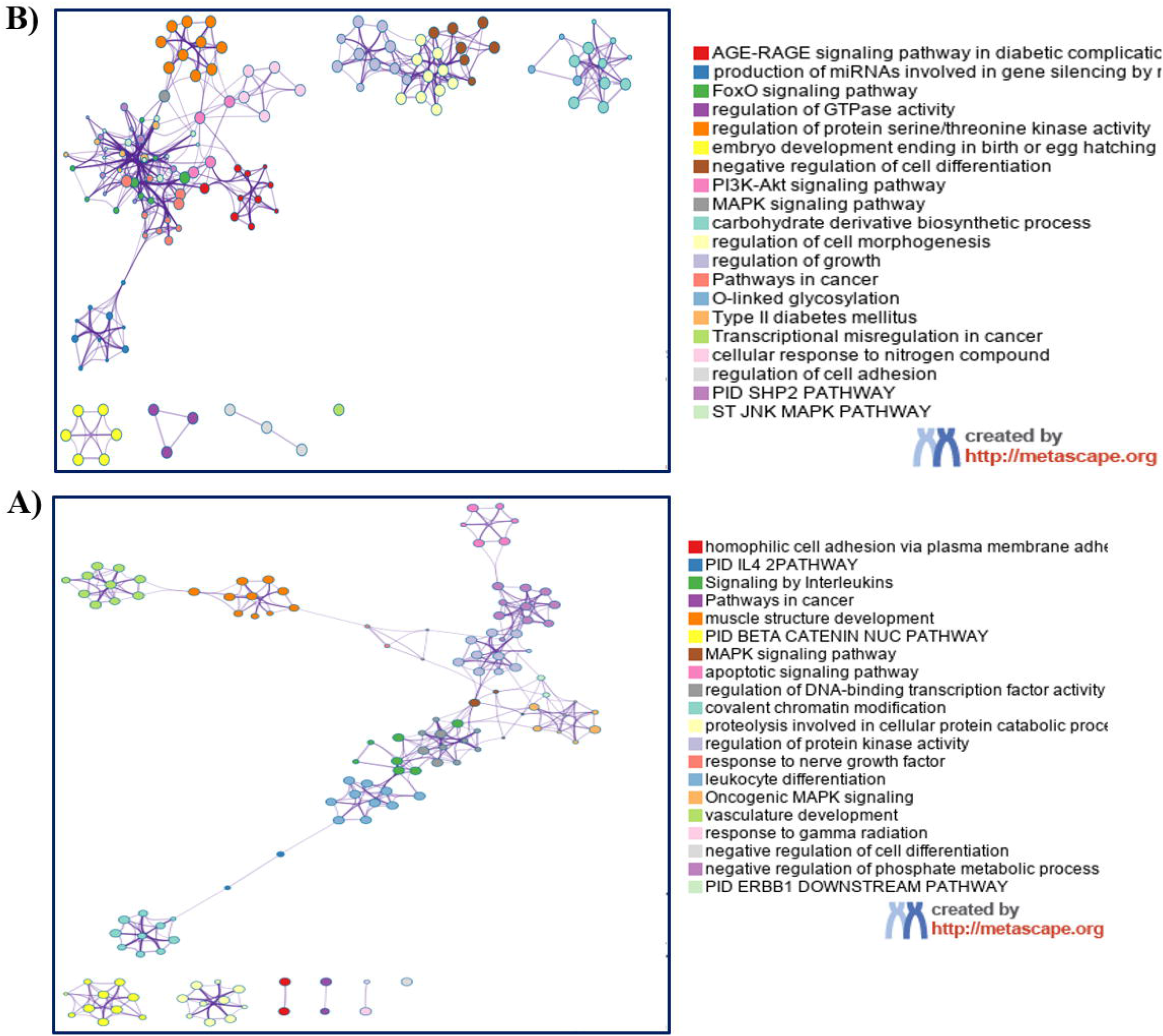
Networks of enriched ontology clusters for Let-7c (A) and miR-155 (B) miRNAs regulated targets. GO: BP, Gene Ontology: Biological Processes terms.

**Figure 3.**
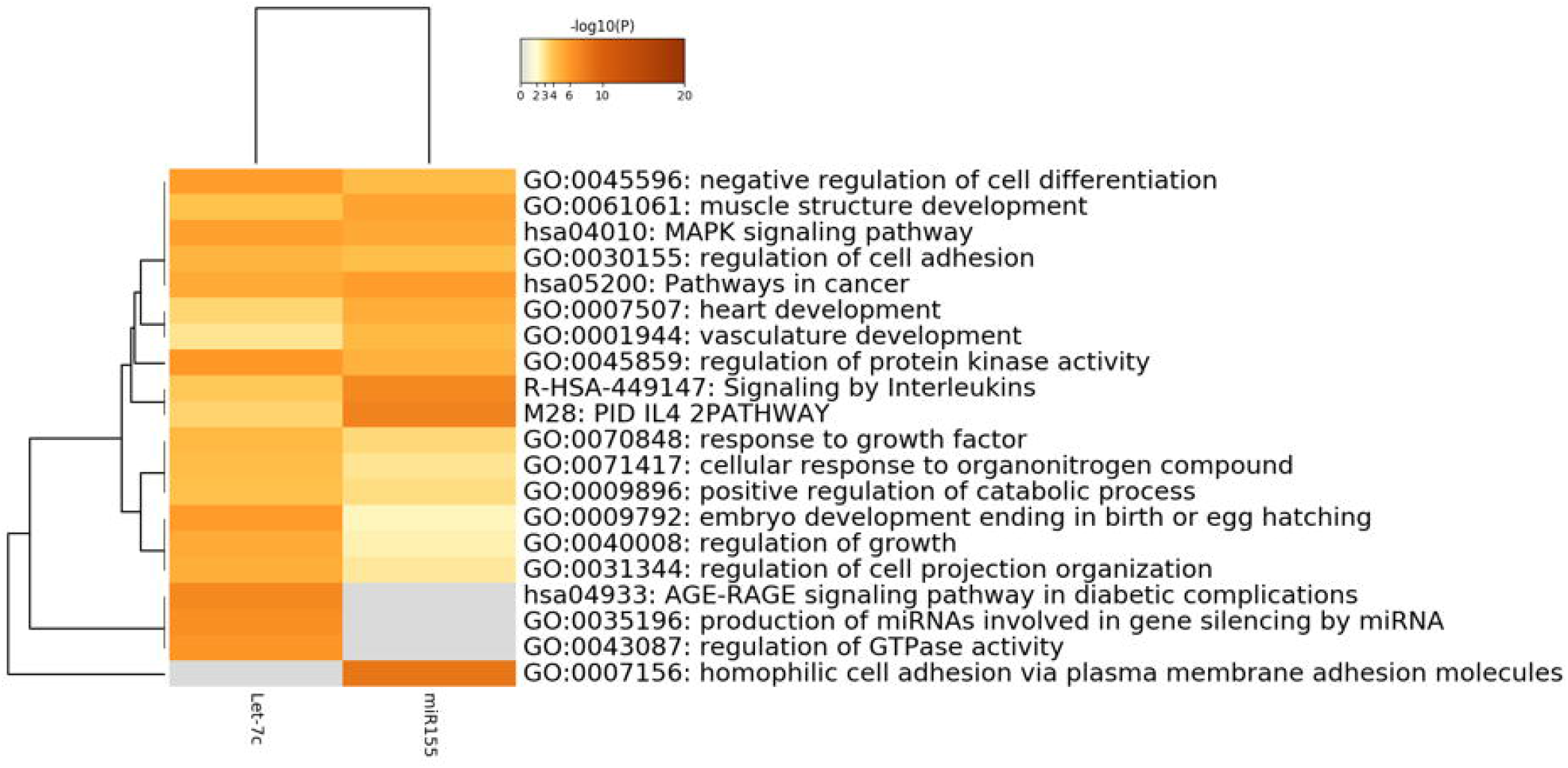
Heatmap of GO: BP enriched terms across miR155 and Let-7c target genes colored by p-values. GO: BP, Gene Ontology: Biological Processes terms.

Bioinformatic analyses of Let-7c gene targets revealed enrichment of signaling pathways associated with diseases, particularly to Type 2 diabetes, such as the AGE/RAGE pathway and the carbohydrate derivate biosynthetic process (Fig 2A). A possible mechanism of Let-7c genetic regulation, is the signaling cascade events of the IGF-1/IGF-1R pathway, which induces the phosphorylation and inhibition of forkhead (FoxO) transcription factors. The FoxO signaling pathway, which is highly enriched among Let-7 targets, is involved in the regulation of both survival and apoptotic genes by inducing the synthesis of the ligand of the apoptosis stimulating fragment (FasL) and the death mediator that interacts with BcL-2 (Bim) (Fig 2A). Overall, these results suggest an essential function for Let-7 in cell survival and carbohydrate metabolism, which may be implicated in the molecular etiology of neurodevelopmental disorders, such as DS.

Among miR-155 unique gene targets, a high enrichment was observed for a group of homophilic adhesion proteins via plasma membrane adhesion composed mostly of clustered alfa protocadherins (Pcdhs) and other nonclustered members (Fig 2B). miR-155 is mainly related to inflammatory pathways such as interleukin signaling. Among the top 20 enriched GO: BP terms for miR-155, no associations with lipid metabolism pathways were found. Searching for lipid-linked pathways, we identified from 100 enriched GO: BP terms one related to long-chain fatty acid import (S2 Fig).

## Discussion

The present study focused on the comparative analyses expression of the miRNAs miR-155 and Let-7c in DS and euploid patients. These miRNAs, located in chromosome 21, have been implicated in neurodegeneration and also have a link to dyslipidemias, and early-stage development of Alzheimer’s pathogenesis.

To improve the therapeutic approach to DS comorbidities, it is necessary to understand them from their molecular level to their clinical features. Consistently with this, a higher difference in the height for DS group compared to the control group was observed. Several reports have found similar differences highlighting short stature as a typical feature in DS, and growth velocity being markedly reduced, mainly between the ages ranging from six months to three years (27). Deficiencies in the secretion of growth hormone (GH) which regulates the production of Insulin-like growth factor (IGFs) have been clearly associated to the marked reduction in speed of growth in young children with DS (28, 29), and consistently with those findings, it has been demonstrated that this phenotype can be normalized with GH therapy (30, 31).

We also observed that BMI was higher in DS compared to the control in the older group analyzed (B groups), while no differences were observed between the younger groups (A control and A DS groups). Previous studies in children and adolescents revealed that children with DS have a healthy BMI, maybe due to parental control in their diet (32). During adulthood, however, there is greater independence of DS individuals for choosing their food, which tends to have a higher caloric content (33). Moreover, risk factors associated with the development of obesity have been described in individuals with DS, such as a decrease in basal metabolism compared to those without DS (34, 35). Additionally, individuals with DS have muscular hypotonia, a phenotype characterized by difficulty in performing activities that involve physical attrition, contributing to a sedentary lifestyle, and, consequently, to weight gain (36, 37).

In addition to dyslipidemias, alterations in the expression of miRNAs could represent other possible risk factors for impaired cognition and a possible link with the early age onset of AD. miRNAs can mobilize through the bloodstream. Therefore, they can be studied by non-invasive methods. The miRNAs Let-7c and miR-155, both present on chromosome 21, were selected for their importance in phenotype variability generating haploinsufficiency of target genes, and their possible relation with the development of Alzheimer’s disease in patients with DS, even at early ages.

We do not observe significative changes for miR-155 expression in peripheral blood from DS people. Previous studies using postmortem brain tissue of 34-42-year-old donors with DS and dementia found the significative up-regulation of miR-155 (9). Interestingly, in the DS mouse model Ts65Dn, miR-155 showed significant overexpression in the hippocampus and the whole blood, but not in lung tissue. Also, there was a down-regulation of the mRNA target Ship1(inositol phosphatase) in the Ts65Dn hippocampus, but not in lung tissue (38). The previous evidence and our results suggest that physiopathological changes in miR-155 expression may be tissue and temporally specific, and it is necessary for an in-depth follow-up study of these changes in the brain tissue to understand its contribution to neurodegeneration in DS.

In murine and cellular models, the role in the development and central nervous system (CNS) functions of several miR-155 gene targets have been demonstrated (9, 39). miR-155 predicted gene target pathways are highly enriched for adhesion molecules such as Pcdhs, which are type I transmembrane receptors expressed predominantly in the central nervous system (CNS) and located in part in synapses involved in pivotal developmental processes, such as axon guidance and dendrite arborization. Changes in their expression might play a role in DS, as has been demonstrated for other neurodevelopmental disorders such as autism, encephalopathy epilepsy, Fragile X syndrome and neurodegenerative diseases (40-42). Ultimately, our results showed a negatively correlation between miR-155 expression and HDL-cholesterol. Although there are not reports for miR-155 mRNA targets involved in lipid metabolism, a deeper *in silico* analysis showed that it may modulate molecules such as fatty acid-binding protein (FABP), which has been used as a cardiac marker because of its function as a long-chain fatty acid carrier in blood and its important role in lipid metabolism (43). Altogether, these observations reinforce its possible role in the development of dementia observed in DS.

Regarding Let-7c expression, the results obtained are consistent with other studies that found a two-fold expression increase in the DS group compared to the euploid population (9, 44). It is also well known that Let-7c is involved in mechanisms related to neurological alterations as an activator of TLR7, in both immune cells and neurons, damaging the CNS through the production of proinflammatory molecules such as the tumor necrosis factor α (TNFα); and the activation of apoptosis via caspases 3, inducing neurodegeneration and neuronal cell death (15). The overexpression of Let-7c into neurons led to morphological and synaptic alterations, and a marked reduction in neuronal excitability through the inhibition of MeCP2 (10), and decreased the expression of the IGF-1R, which is expressed differentially throughout the brain (45). Furthermore, IGF-1R/Insulin alterations are characteristic in patients with AD, Parkinson’s, and other neurodegenerative disorders. IGF-1 is an essential factor for healthy growth and development, involved in neuronal survival, myelin synthesis, cell metabolism, astrocyte function, angiogenesis, neuronal excitability, and oligodendrogenesis (46). A decrease in IGF-1 has also been reported in people with DS (29, 47, 48). Moreover, the overexpression of Let-7c in the brain tissue of DS adults with dementia was observed, demonstrating its involvement with this process (9). Let-7c overexpression is also related to mitochondrial cardiomyopathies. In trisomic hearts, down-regulation was observed of the SLC25A4/ANT1 gene (Solute carrier family 25 member 4/ Adenine nucleotide translocator 1), a predicted target of Let-7c and associated to mitochondrial and cardiac anomalies (14). Also, mitochondrial dysfunction is closely linked to the increase in oxidative stress, which has a central role in the pathogenesis of DS and the etiology of different intellectual disabilities, as it affects highly oxidative tissues such as the brain, suggesting a synergism between Let-7c and oxidative stress (49-52).

Integrating alterations in miRNA expression with dyslipidemias for the physiopathology in DS neurodegeneration, we start from the evidence that indicates dyslipidemias are a risk factor for the development of dementias such as Alzheimer’s (53-55). Elevated cholesterol levels induce accelerated APP processing in its amyloidogenic pathway, a condition related to the development of Alzheimer’s dementia (56-59). Studies of the lipid profile in children with DS, compared with their euploid siblings, adjusting for BMI showed that even in normal body weight, the DS children presented alterations in cholesterol, triglycerides, HDL, and LDL (16, 19). The results obtained in this study demonstrate that individuals with DS have alterations in the lipid profile compared to controls at an early age as an increase in cholesterol, triglycerides and low levels of HDL; and consistent in adults with DS. These results support the idea that the alterations in lipid metabolism are not only due to obesity, but possibly the presence of an extra copy of chromosome 21; meaning a risk factor for AD neurodegeneration. Several reports have determined a positive correlation of Aβ levels with elevated cholesterol and low HDL levels (55, 60, 61). HDL maintains the transport of cholesterol to neurons for functioning and has a role in Aβ clearance; hence, high cholesterol levels and low HDL levels are related to cognitive disorders (55, 60-62). In line with this evidence, we found that Let-7c overexpression was higher among DS individuals with low-HDL levels and that miR-155 levels negatively correlate to low HDL levels. Even though not mRNA targets for miR-155 and Let-7c involved in lipid metabolism have been demonstrated, and based on the *in-silico* analysis, a direct relationship between their expression and alterations in the lipid profile cannot be discarded. However, the fact that there is a negative correlation of their expression with the concentration of HDL or a differential expression according to lipid levels suggests the existence of a molecular network linking both parameters.

Overall, these results suggest that a set of differentially expressed miRNAs promotes the progression of cognitive impairment in patients with DS by regulating genes of biological pathways involved in lipid metabolism and nervous system development. Moreover, other biological processes such as inflammation and the immune system must integrate for Alzheimer’s neurodegeneration in the DS puzzle.

## Conclusion

Altogether, our results reveal a pattern of miR-155 and Let-7c miRNAs expression that correlates with the alteration in the concentration of HDL, reinforcing the hypothesis of the participation of this lipoprotein in the homeostasis of molecules related to cognitive function and early development of dementias, a frequent feature in Down syndrome. The data suggests that in Down syndrome, Let-7c and miR-155, located on the triplicated chromosome 21, maybe dysregulated and playing a key role in the pathogenesis of Alzheimer’s dementia. This implies that modulating the expression of these miRNAs in the CNS may help abrogate the inevitable pathway to dementia in adults with Down syndrome.

## Data Availability

We declare that all data referred to in the manuscript is available and subject to any revision.

## Acknowledgments

Carla Angulo-Rojo and Javier Magaña Gómez received financial support from Universidad Autónoma de Sinaloa (PROFAPI2014/015 and PROFAPI2015/015). Jesús Manuel Pérez-Villareal received a Master fellowship from CONACyT (CVU785590). Katia Aviña-Padilla received a postdoctoral fellowship from DGAPA-UNAM and acknowledges CABANA program for training in bioinformatics. Technical support for this work was obtained from Luis Alberto Aguilar Bautista, Alejandro de León Cuevas, Carlos Sair Flores Bautista and Jair García of the Laboratorio Nacional de Visualización Científica Avanzada (LAVIS).

## Supporting information

**S1 Fig. Circous graphs of overlapping and divergent targets genes of miRNA 155 and Let-7c**. A) only at the gene level, purple curves link identical genes; B) including the shared term level, blue curves link genes that belong to the same enriched ontology term between the targets of both miRNAs. The inner circle represents gene lists, where hits are arranged along the arc. Genes that hit multiple lists are colored in dark orange, and genes unique to a list are shown in light orange.

**S2 Fig. Networks of enriched ontology clusters for miR-155 miRNA regulated targets**. 100 GO: BP, Gene Ontology: Biological Processes terms.

## Notes

### Competing Interest Statement

The authors have declared no competing interest.

### Funding Statement

CAR obtained financial support for this project from Universidad Autonoma de Sinaloa
(PROFAPI2014/015 and PROFAPI2015/015) (https://www.uas.edu.mx/). KAP received
a Postdoctoral fellowship from Universidad Autonoma Nacional de Mexico (DGAPAUNAM)
(https://www.unam.mx/) and JMPV received a Master fellowship from Consejo Nacional de Ciencia y Tecnologia (CONACyT) (https://www.conacyt.gob.mx/). We declare that sponsors did not play any role in the study design, data collection, analysis, decision to publish, or preparation of the manuscript.

### Author Declarations

The study followed the provisions of the Declaration of Helsinki and Good Clinical Practice guidelines, and the local institutional review board approved the protocol. The research project was reviewed and approved by the Research Ethics Committee at the Medicine Faculty of Autonomous University of Sinaloa, which has a national certification (CONBIOETICA-25-CEI-003-20181012) for its duties. Parents or legal guardians of people with DS were informed before the evaluation and provided written consent to undergo study procedures.

