## Supplementary figures and images for "Profiling of circulating chromosome 21-encoded microRNAs, miR-155 and Let-7c, in Down Syndrome People"

### S1 Fig.tif

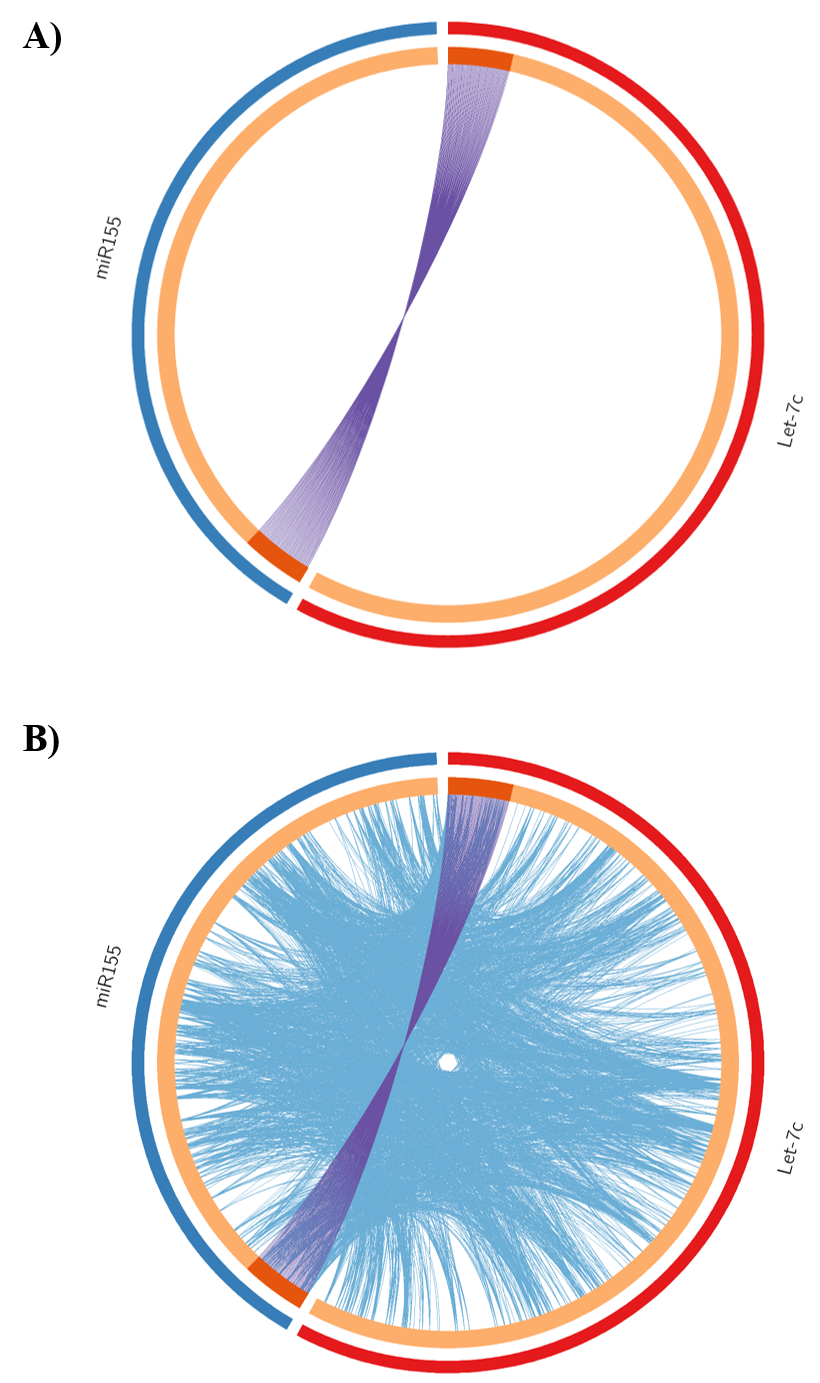

### S2 Fig.tif

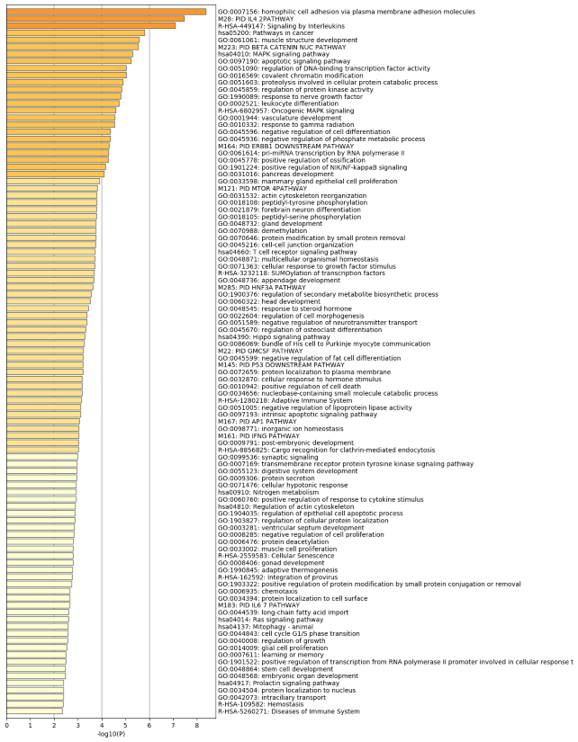
